# Attentional compensation in neurodegenerative diseases: the model of premanifest Huntington’s disease mutation carriers

**DOI:** 10.1101/2020.06.03.20121079

**Authors:** Lorna Le Stanc, Marine Lunven, Maria Giavazzi, Agnès Sliwinski, Pierre Brugières, Katia Youssov, Anne-Catherine Bachoud-Lévi, Charlotte Jacquemot

## Abstract

The ability of the brain to actively cope with neuropathological insults is known as neural compensation. It explains the delayed appearance of cognitive symptoms in neurodegenerative diseases. In contrast to the neural signature of compensation, its cognitive counterpart is largely unknown due to the difficulty of identifying cognitive dysfunctions concealed by compensation mechanisms. We combined computational modelling and neuroanatomical analysis to explore cognitive compensation. We used Huntington’s disease (HD) as a genetic model of neurodegenerative disease allowing to study compensation in premanifest mutation carriers (preHDs) free from overt cognitive deficits despite incipient brain atrophy.

Twenty preHDs, 28 HD patients and 45 controls performed a discrimination task. We investigated the processes underlying cognitive compensation using drift diffusion models. They assume that the discrimination process relies on the accumulation of evidence at a certain rate and terminates when a response threshold is reached.

HD patients’ performances were lower than controls’ and explained by a higher response threshold and a lower accumulation rate compared to controls. PreHDs performed similarly to controls but had a response threshold between those of controls and HD patients. This nascent increase in response threshold predicted the accumulation rate, which was faster than controls. This suggests that the higher accumulation rate conceals the nascent deficit in response threshold corroborating the capacity of the brain to resist neuropathological insults in preHDs. The higher accumulation rate was associated with parietal hypertrophy in mutation carriers, and with higher hippocampal volumes in preHDs suggesting that cognitive compensation may rely on attentional capacities.

**Significance statement:** Enhancing mechanisms compensating brain degeneration in neurodegenerative diseases might allow to delay their onset and progression. Yet, the cognitive mechanisms of compensation remain to be identified. In order to explore this issue, we used Huntington’s disease as a genetic model of neurodegenerative diseases and combined computational modelling (drift diffusion models) and neuroanatomical data analysis. In the early stage of the disease, before the appearance of overt cognitive symptoms, we showed the involvement of the left superior parietal cortex and hippocampus in maintaining normal behavioural performances. This suggests that attention is used to compensate for brain atrophy early in the disease. This work describes promising means of measuring and understanding compensation mechanisms in neurodegenerative diseases and might help developing new therapies.

## Introduction

Brain atrophy precedes intellectual deterioration in neurodegenerative diseases (Tabrizi et al., 2009). Normal behaviour is maintained until the pathological load becomes too great, leading to the appearance of clinical symptoms (Papoutsi et al., 2014; Gregory et al., 2018; Soloveva et al., 2018). The ability of the brain to actively cope with neuropathological insults underlies the cognitive reserve, which depends on lifetime intellectual activities and environmental factors. It is based on the brain’s capacity to increase the efficiency of an existing yet deteriorating network (neural reserve) and/or to recruit other regions when performing a task (neural compensation) (Barulli and Stern, 2013; Soloveva et al., 2018). Neurodegenerative diseases do not affect all brain parts and functions equally and simultaneously; some cognitive functions may compensate for others impacted earlier. Whereas the concept of compensation is widely accepted (Papoutsi et al., 2014; Gregory et al., 2018), the cognitive functions underlying it are still unknown mainly because methods for studying and measuring them are lacking.

Assessing cognitive dysfunction concealed by cognitive compensation requires the identification and the disentanglement of underlying cognitive dysfunction and compensation mechanisms. Generally, neurodegenerative diseases remain undiagnosed until disease manifestation – a point at which it is difficult to study cognitive compensation since compensation mechanisms are no longer effective. Huntington’s disease is an inherited, monogenetic (expanded CAG repeat in the huntingtin gene), dominant, and fully penetrant neurodegenerative disease (Tabrizi et al., 2011). Individuals with more than 40 CAG repeats will develop the disease. This allows for identifying premanifest Huntington’s disease gene carriers (preHDs) before the clinical onset of the disease and motor, cognitive and psychiatric deterioration (Ross et al., 2019). Therefore, Huntington’s disease is a particularly well-suited neurodegenerative model for studying compensation mechanisms from genetic diagnosis to onset of overt clinical manifestations (Tabrizi et al., 2011; Malejko et al., 2014; Gregory et al., 2018).

In this study, we used drift diffusion models (DDMs) (Ratcliff and Mckoon, 2008; Mulder et al., 2014) to decipher cognitive dysfunction from compensation mechanisms that we would otherwise be unable to study separately (Wiecki et al., 2016; Zhang et al., 2016; Anders et al., 2017). DDMs unravel the different steps of the decision process when having to choose between two alternatives (A or B). They rely on the hypothesis that one needs to accumulate a certain amount of sensory evidence in order to decide between the two alternatives. The amount of evidence accumulated and its rate of accumulation are obtained by fitting the distributions of responses (alternative A or B) and response time combined.

We applied DDMs to preHDs without overt cognitive symptoms, patients at an early stage of Huntington’s disease (earlyHDs), and controls. PreHDs perform at least as well as healthy participants in most cognitive tasks, despite displaying incipient atrophy of the striatum (Snowden et al., 2002; Tabrizi et al., 2009; Stout et al., 2012) and functional changes (Feigin et al., 2006; Klöppel et al., 2009; Wolf et al., 2012) suggesting the deployment of compensation mechanisms in most cognitive domains. Yet, language impairments have been reported in preHDs in small cohorts of participants (de Diego Balaguer et al., 2008; Németh et al., 2012; Hinzen et al., 2018). We therefore designed a language discrimination task in which participants were asked to decide whether two pseudowords were identical or different. This task relies on an automatic linguistic process that does not require any learning procedure (Näätänen et al., 1997; Dehaene-Lambertz and Baillet, 1998). We first analysed behavioural data, assuming that preHDs would not show overt deficits whereas earlyHDs should have some, assuming that their compensation mechanisms would no longer be effective. We then used DDMs to identify subclinical deficits and cognitive compensation mechanisms in preHDs. Finally, in order to explore the correlation between cognitive compensation mechanisms and brain structure, we assessed the neuroanatomical correlates of these mechanisms.

## Materials and methods

### Participants and clinical assessment

We recruited 93 native French-speaking participants between December 2013 and July 2017. Forty-eight were Huntington’s disease mutation carriers evaluated with the Unified Huntington’s Disease Rating Scale (Huntington Study Group, 1996), the Mattis Dementia Rating Scale (Mattis, 1976), forward digit span and categorical fluency. Twenty-eight were at an early stage of the disease (earlyHDs; stages I and II of the classification based on the total functional capacity score of the Unified Huntington’s Disease Rating Scale), and the other 20 were at the premanifest stage (preHDs; total functional capacity score of 13 and total motor score below five (Tabrizi et al., 2009) and no overt cognitive deficits (Ross et al., 2019)). Forty-five were healthy participants recruited as controls and matched with the earlyHDs and preHDs for demographic variables, such as sex, handedness, years of education, and age (all *p*>0.05). The two mutation carrier groups were also matched for demographic variables (all *p*>0.05) except for age (*p*<0.05). The participants had no neurological or psychiatric disorders other than Huntington’s disease in the mutation carriers. The demographic and clinical description of participants are summarised Table 1 and Table 2.

**Table 1.**
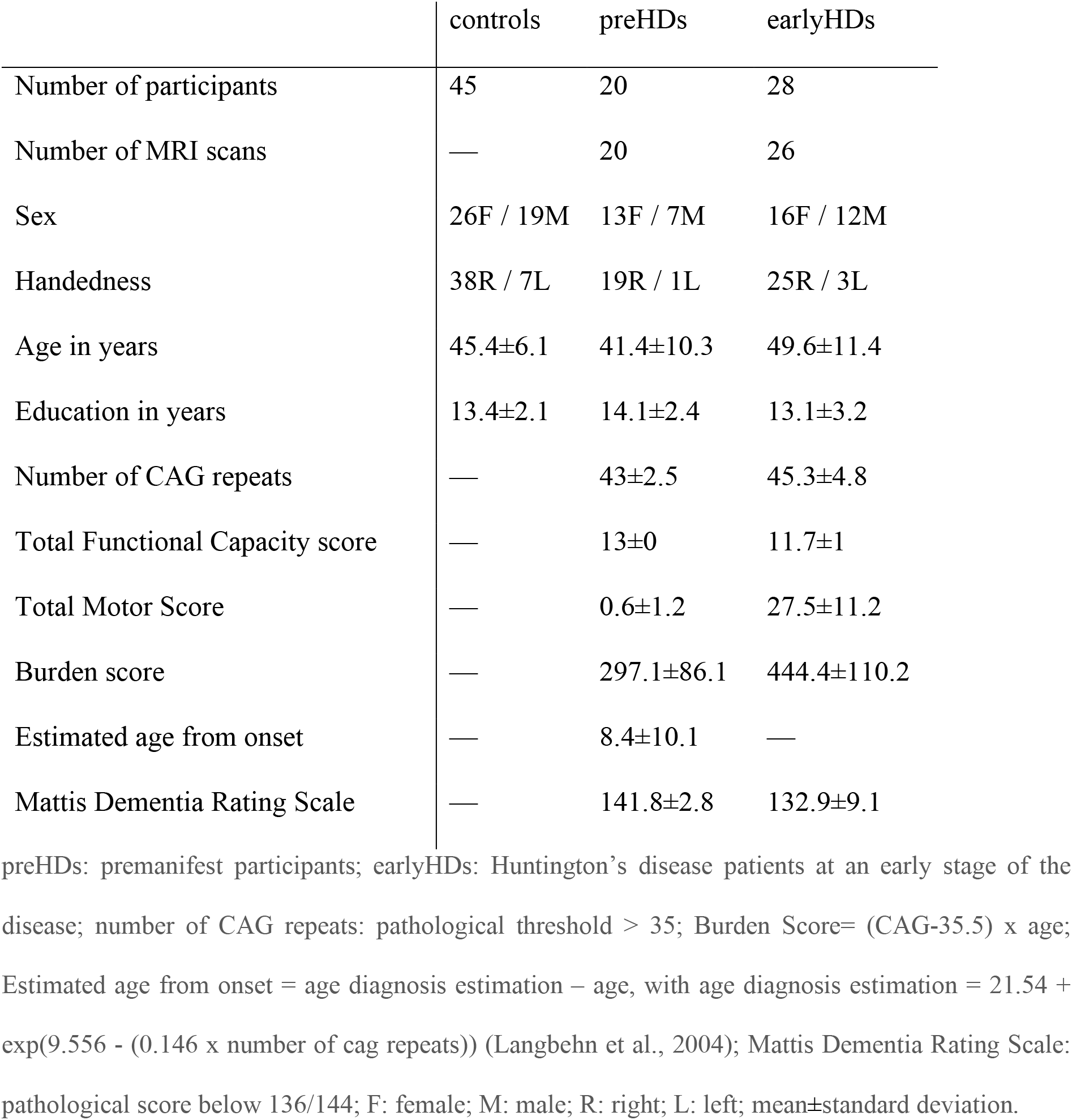
Demographic and clinical data for participants

**Table 2.** Cognitive assessment

|  | Descriptive statistics |  |  | Group comparisons |  |  |
| --- | --- | --- | --- | --- | --- | --- |
|  | controls* | preHDs | earlyHDs | preHDs/controls | earlyHDs/controls | preHDs/earlyHDs |
| Forward digit span | 6.4±1 | 6.5±1.3 | 5.6±1 | [-0.7,0.7], $p=0.99$ | [-1.4,-0.1],<br>$p=0.016$ | [0.01,1.5],<br>$p=0.0454$ |
| Categorical fluency | 33.8±8.1 | 37.6±7.1 | 21.8±9.1 | [-2.6,9.5], $p=0.37$ | [-16.7,-5.5],<br>$p<0.001$ | [8.5,20.7],<br>$p<0.001$ |
| Verbal fluency | 75.6±22.8 | 68.3±22.9 | 36.5±16 | [-23.1,7.8],<br>$p=0.47$ | [-51.0,-23.1],<br>$p<0.001$ | [13.8,45.0],<br>$p<0.001$ |
| Stroop colour | 83.6±11.2 | 75.7±14 | 48.6±13.7 | [-18.5,1.0],<br>$p=0.09$ | [-38.9,-21.6],<br>$p<0.001$ | [11.9,31.3],<br>$p<0.001$ |
| Stroop interference | 47.5±10.5 | 47.2±10.2 | 29.9±12 | [-8.9,7.0], $p=0.96$ | [-21.0,-7.0],<br>$p<0.001$ | [5.2,20.9],<br>$p<0.001$ |
| Stroop word | 104.7±16.8 | 98.4±15.8 | 66.7±12.3 | [-18.4,4.8],<br>$p=0.35$ | [-45.3,-24.7],<br>$p<0.001$ | [16.7,39.7],<br>$p<0.001$ |
| Symbol digit<br>modalities Test | 49.5±7.7 | 54±11.1 | 28.7±7.8 | [-3.5,9.9], $p=0.50$ | [-23.7,-11.4],<br>$p<0.001$ | [14.1,27.3],<br>$p<0.001$ |

Forward digit span, categorical fluency and Unified Huntington's Disease Rating scale cognitive scores.
Descriptive statistics report mean±standard deviation for each group. Group comparisons report Tukey's
post-hoc test [95% confidence interval], $p$ -value for each group comparison when there was a main
effect of group. preHDs: premanifest participants; earlyHDs: Huntington's disease patients at an early
stage of the disease; \* Cohort of controls restricted to 27 participants.

This study was performed in accordance with the Declaration of Helsinki (2008). Participants were recruited from a clinical biomarker study (NCT01412125) in outpatients approved by the ethics committee of Henri Mondor Hospital (Créteil, France). Participant inclusion ended when 45 valid brain MRI scans had been obtained from mutation carriers. All participants gave written informed consent and were tested at Henri Mondor Hospital or at the Ecole normale supérieure (Paris, France).

### Experimental design

We designed a language discrimination task, using pseudowords, in which participants had to determine whether two pseudowords were identical or different (Fig. 1A).

**Figure 1.**
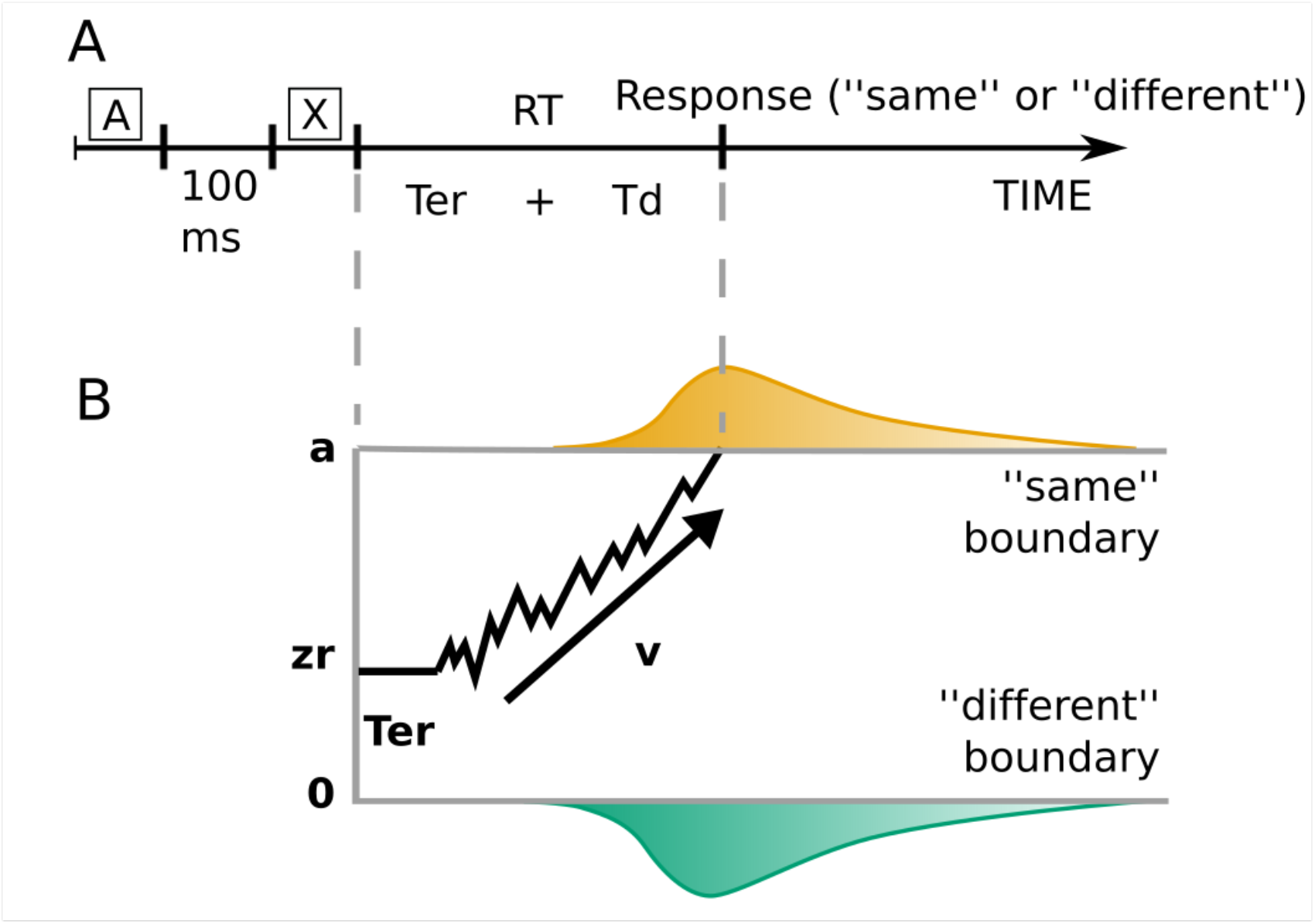
Discrimination task and hierarchical drift diffusion model. (A) Participants heard two pseudowords (A and X) separated by the 100 ms and had to decide whether they were identical or not. The response time (RT) is the sum of the non-decision time (Ter) and the decision processes (Td): RT=Ter+Td. (B) Example of the trajectory of the drift diffusion model for a “same” trial in which the correct response was delivered. Two decision boundaries (0 and a) represent the “same” and “different” decisions. The drift rate, v, represents the rate of evidence accumulation. The diffusion process starts between the two boundaries at zr (= 0.5 if not biased toward one of the alternatives) and continues until it reaches one of the two boundaries. The predicted response time is the sum of the durations of the diffusion process called decision time and the non-decision time encompassing stimulus pre-processing and motor planning and execution.

The pseudowords had one to three syllables and zero to two consonant clusters. Items consisted of a pair of pseudowords presented 100 ms apart. The two pseudowords were identical in half the items and differed by a single consonant (e.g. /tiplysk/ and /tipʁysk/) in the other half. The location of the consonant that differed was varied between trials to prevent expectation. Twelve items were presented per number of syllables (one to three), consonant cluster (0 to two) and type of trial (“same/different”), resulting in 216 trials in total. A female native French speaker (the last author) pronounced the pseudowords for the recording, with each pseudoword lasting 1030+165 ms.

The participants sat in a quiet room, in front of an Apple MacBook Pro (AZERTY keyboard), wearing headphones adjusted for hearing comfort. They were asked to press the “s” key labelled “P” for “pareil” (same) if the two sequences were identical, or the “l” key labelled “D” for “différent” (different) if they differed. The experiment began with a training session of four trials with feedback, followed by the experimental trials without feedback. Response (“same/different”), accuracy (correct or incorrect response) and response time were recorded after each item. The response triggered the presentation of the next trial, 1000 ms later. The task lasted less than 10 minutes in total. Except for the training session, trials were randomized within two blocks separated by a break.

### Statistical analyses of behavioural data

#### Clinical cognitive assessment

We analysed the effect of group on the forward digit span, categorical fluency and Unified Huntington’s Disease Rating Scale cognitive scores using ANOVAs with group as a between-participants factor and age as a covariate to control for the difference in age between the mutation carrier groups.

#### Analyses of response times and accuracy

The training trials were not included in the analyses. Trials in which participants withdrew temporarily from the experiment and answered before the end of the trial item were removed, resulting in a 0.06% loss of data. Accuracy analyses were run on the remaining trials after calculating the mean accuracy by subject. Response time analyses were run on correct trials (3.9% loss) lasting more than 150 ms (8.1% loss) and after logarithmic transformation to ensure a normal distribution.

We analysed the effect of group on mean accuracy using a linear model and response time using a linear mixed effects model (models that can deal with unbalanced data sets and missing data contrary to ANOVAs). We focused on three group comparisons of interest: preHDs/controls, earlyHDs/controls and preHDs/earlyHDs using the package “multicomp” with single-step correction from R version 3.4.0. We included age as a covariate in both analyses. In the response time analyses, the maximum random structure allowing convergence without over fitting included participant and item as random intercepts.

#### Model fit and selection

We used DDMs (Ratcliff and Mckoon, 2008; Mulder et al., 2014) to analyse the mechanisms underlying the forced decision between “same” and “different”. These models assume that sensory evidence is accumulated at a certain speed, called the drift rate (v), up to a response threshold (a) triggering the motor response. Accumulating evidence takes time. It is a noisy process requiring multiple evidence samples to extract information from the stimulus before enough evidence is collected to make a decision. The time required for non-decisional processes, such as stimulus processing, motor preparation and execution, is captured in the non-decision time (Ter). The *a priori* bias towards one of the alternatives is called the relative bias (zr) (Fig. 1 B). These four parameters (v, a, Ter and zr) are obtained by fitting the distribution of responses (“same” or “different”) and the corresponding response time at each trial. In our task, the participant hears two pseudowords. First, the acoustic information is transformed into neural information (stimulus processing). Then, the phonological distance between them is assessed in the brain at a certain speed (drift rate of evidence accumulation). According the participant’s conservatism, the amount of evidence required to decide will be more or less elevated (response threshold). Once the decision is taken, the participant presses the corresponding key (motor preparation and execution).

We used Bayesian hierarchical DDMs (Wiecki et al., 2013), currently the most efficient method for dealing with small numbers of observations (Ratcliff and Childers, 2015). This approach assumes that individual parameter estimates are random samples of group-level distributions. It provides probability distributions for parameters, called posterior distributions, rather than single-value estimates. Data were cleaned as in behavioural analyses, but Bayesian hierarchical DDMs use both correct and incorrect responses and response times without logarithmic transformation. We assumed the same absolute drift rate value for both answers (“same” and “different”), allowing for a possible relative bias.

We tested two variants of the Bayesian hierarchical DDMs, with different group-level parameters.

In the first model (full model), each parameter had three group-level distributions, corresponding to the three groups of participants (controls, preHDs, earlyHDs). The second model (parsimonious model), assumed that only the response threshold and drift rate had different group-level distributions. Using the recommended procedures to fit and assess model convergence (Wiecki et al., 2013) (Supplementary Methods 1), we selected the parsimonious model; as the full one did not capture any additional data patterns (Supplementary Methods 2 and Supplementary Table 1).

#### Analysis of model parameters

We analysed the effect of group on the parameters with group-level distributions: response threshold and drift rate. The hierarchical structure of the model violates the independence assumption of classical frequentist statistics. Bayesian statistics were used for direct comparisons of group posterior distributions. Bayesian probabilities are denoted *P(hypothesis)* and express the probability of a hypothesis being true. As a mock example, we can test the hypothesis that earlyHDs have a higher drift rate than controls *(P(v_eartyHDs_>v_controis_))*. A probability of 0.95 would indicate that there is 95% chance that it is true, while a probability of 0.05% would indicate that there is 95% chance that earlyHDs have instead a lower drift rate compared to controls *(P(v_eartyHDs_>v_controls_)* = *1*-*P(v_earlyHDs_<v_controls_))*. A probability of 0.5 would indicate that both hypotheses (higher or lower drift rate) are equally probable.

#### Statistical analysis of structural imaging data

Forty-six brain MRI scans were obtained within about three months of behavioural data acquisition in gene carriers (20 preHDs and 26 earlyHDs). They were compared with 30 scans from external healthy participants (imaging controls), matched with the mutation carriers for age and sex (46.1 ± 13.9 years old, 15 females). We first studied the differences in subcortical and cortical structure between the mutation carriers and imaging control groups. We then studied the relationship between brain structure and the behavioural measures (response time, accuracy, drift rate and response threshold) for the mutation carriers.

#### MRI acquisition and preprocessing

Three-dimensional T1-weighted structural scans were acquired with a MP-RAGE sequence on a Siemens symphony 1.5 Tesla whole-body scanner (Henri Mondor Hospital, Paris, France) with a 12-channel head coil (TR=2400 ms, TE=3.72 ms, TI=1000 ms, FA=8°, FOV=256*256 mm^2^, 1-mm isotropic voxel, slice thickness=1 mm, no inter-slice gap, 160 sagittal sections).

MRI scans were preprocessed with Freesurfer (http://surfer.nmr.mgh.harvard.edu/) (Fischl et al., 2002). The procedure included the removal of non-brain tissue, normalization of the intensity of the grey/white matter boundary, automated topology correction, and surface deformation. The following subcortical structures were automatically segmented: thalamus, caudate, putamen, pallidum, hippocampus, amygdala and the nucleus accumbens. Cortical thickness (in mm) was calculated as the shortest distance between the grey/white matter boundary and the pial surface at each vertex across the cortical mantle (Fischl and Dale, 2000). All reconstructed data were visually checked for segmentation accuracy by a neuropsychologist (ML) trained to brain structural segmentation analysis and reviewed by an expert neurologist blinded to participants genetical status. The spherical cortical thickness data for all subjects were mapped onto an “average” subject by surface-based registration methods (Fischl et al., 1999), to match morphologically homologous cortical locations between subjects. We used a 10 mm full-width at half-maximum Gaussian kernel to smooth maps of cortical thickness.

#### Neuroanatomical differences between groups

We analysed the neuroanatomical differences between imaging controls, preHDs and earlyHDs.

For each subcortical volume normalized to the total intracranial volume, we performed an ANOVA, with group as a main effect and age as covariate. ANOVA’s *p*-values were divided by seven to correct for multiple comparisons on the seven subcortical structures (Bonferroni correction).

In the cortex, vertex-wise comparisons of cortical thickness values between groups were performed in Freesurfer with generalised linear models, with cortical thickness as the dependent variable, group as the predictive factor, and age as covariates. At each vertex, F-statistics were calculated to test the hypothesis of a difference in cortical thickness for each group comparison (two-tailed test). We corrected for multiple comparisons by family-wise error cluster-based correction, using Monte Carlo simulations with 10,000 iterations.

#### Relationship between brain structure and behavioural measures in mutation carriers

We explored the relationship between the neuroanatomical structure and behavioural measures (mean accuracy, mean response time, drift rate and response threshold) in mutation carriers depending on the disease stage (preHDs or earlyHDs).

For each subcortical structure/measure combination, we fitted a linear model with the measure as the dependant variable, the subcortical volume normalized to the intracranial volume and disease stage as predictive variables, and age as covariate. We tested the interactions between disease stage and the subcortical volume. *P*-values were Bonferroni-corrected for multiple comparisons over the seven subcortical structures. If the interaction was not significant, the disease stage was removed from the analysis before testing the effect of the volume on the given measure.

In the cortex, we fitted one generalised linear model for each measure with the cortical thickness as the dependant variable, the measure and the disease stage as predictive variables, and age as covariates. At each vertex, *F*-statistics were calculated to test the hypothesis of an interaction between cortical thickness and disease stage. We corrected for multiple comparisons by family-wise error cluster-based correction, using Monte Carlo simulations with 10,000 iterations. If there was no cluster with a significant interaction, the disease stage was removed from the analysis before testing the hypothesis of the non-null relationship (two-tailed test) between the measure and the cortical thickness. The analyses described being conservative, we performed one-tailed test (*t*-statistics) testing the hypothesis of a positive or negative relationship. We report the results in the supplementary material (Supplementary Fig. 1 and Supplementary Table 2).

Finally, in order to locate preHDs and earlyHDs compared to healthy participants in the significant clusters, we used the clusters identified by the generalised linear model analyses as regions of interest, from which we extracted cortical thickness values for imaging controls and mutation carriers. We tested for differences in cortical thickness between preHDs and imaging controls and earlyHDs and imaging controls by performing an ANOVA on the mean cortical thickness for each significant cluster.

## Results

### Analysis of behavioural data

#### Clinical cognitive assessment

ANOVAs showed significant main effects of group (controls, preHDs, earlyHDs) for the forward digit span, the categorical fluency and each cognitive scores of the Unified Huntington’s Disease Rating Scale. Tukey’s post-hoc analyses revealed that the earlyHDs were impaired on each test compared to preHDs and controls (all *p*<0.05), whereas preHDs performances were similar to controls (all *p*>0.05) (Table 2).

#### Analysis of response times and accuracy

EarlyHDs were less accurate and slower than controls and preHDs ([earlyHDs/controls] accuracy: *β*=-0.011±0.002, 95%CI=[-0.015,-0.007], *t*=-6.8, *p*<0.001; response time: *β*=0.060±0.007, 95%CI=[0.044,0.076], *z*=8.7, *p*<0.001. [earlyHDs/preHDs] accuracy: *β=*-0.006±0.001, 95%CI=[-0.008,-0.004], *t*=-5.9, *p*<0.001; response time: *β*=0.032±0.004, 95%CI=[0.022,0.042], *z*=7.5, *p*<0.001). By contrast, preHDs performed as well as controls (accuracy: *β=*0 001±0.002, 95%CI=[-0.004,0.005], *t*=0.3, *p*=0.97; response time: *β=*-0.003±0.008, 95%CI=[-0.021,0.015], *z*=-0.4, *p*=0.93) (Fig. 2A-B).

**Figure 2.**
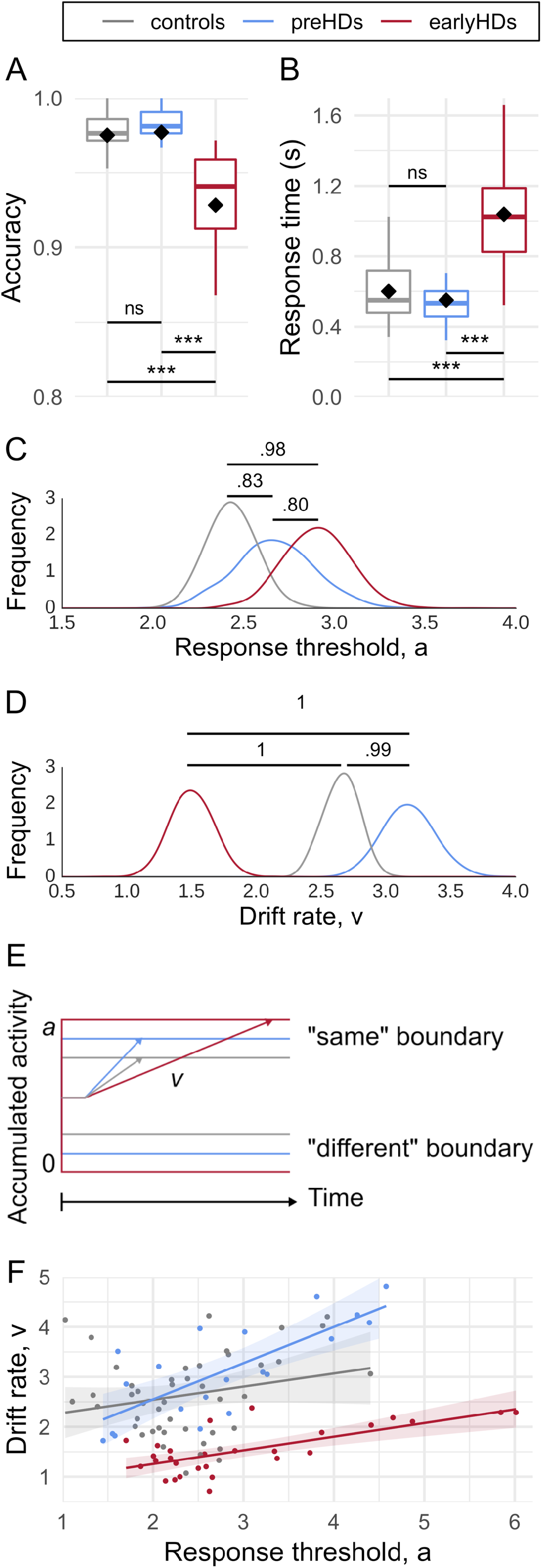
Results of behavioural data analyses. Boxplots of (A) Accuracy and (B) Response time (s) for each group. In boxplots, the middle hinge corresponds to the median, the lower and upper hinges correspond respectively to the first and third quartiles. The whiskers extend from the hinge to the largest value no further than 1.5*inter-quartile range of the hinge. Diamonds represent the means, ***: *p*<0.001, ns: non-significant. (C) Group probability distribution (posterior estimates) of the response threshold, a, and (D) drift rate, v. Bayesian probabilities are reported. (E) Schematic representation of model parameters for each group. (F) Relationship between the drift rate (y-axis) and the response threshold (x-axis). Points represent individual values, lines and shades around them represent the linear fit and the confidence interval. Controls are represented in grey, premanifest participants (preHDs) in blue and Huntington’s disease patients at an early stage of the disease (earlyHDs) in red.

#### Analysis of model parameters

The group probability distribution (posterior estimates) of response threshold and drift rate are displayed in Figure 2C and 2D. The results are schematically represented Figure 2E. The earlyHDs had a higher response threshold *(P(a_earlyHDs_>a_controls_)*=0.98) and a lower drift rate than controls *(P(v_earlyHDs_<v_controls_)*=1). EarlyHDs therefore needed to accumulate more evidence to discriminate between “same” and “different”, and they accumulated it slower than the other groups.

The response threshold for preHDs was between earlyHDs’ *(P(a_preHDs_<a_earlyHDs_)*=0.80) and controls’ *(P(a_preHDs_>a_controls_)*=0.83). PreHDs’ drift rate was higher than both controls *(P(v_preHDs_>v_controls_)*=0.99) and earlyHDs *(P(v_preHDs_>v_earlyHDs_*)=1), indicating faster evidence accumulation.

Post-hoc, we fit a Bayesian linear model (Goodrich et al., 2020) to predict the drift rate based on the response threshold and the group. Age was used as a covariate. The effect of response threshold on the drift rate was positive (*β*=0.29, *P*(*β*>0)=0.98) and similar for earlyHDs and controls *(P(β_earlyHDs_> β_controls_)*=0.46). The effect of response threshold was stronger in preHDs *(β_preHDs_*=0.16, *P(β_preHDs_>*0)=1, interaction *P(β_preHDs_> β_controls_)*=0.99). This indicates that an increase in response threshold predicted an increase in drift rate which is similar in controls and earlyHDs but stronger in preHDs (Fig. 2F).

### Analysis of structural imaging data

#### Neuroanatomical differences between groups

There was a main effect of group on the volumes of caudate, putamen, accumbens, pallidum and thalamus (all *p*-values<0.05) (Fig. 3A). On these structures, Tukey’s post-hoc analyses revealed that earlyHDs had a lower grey matter volume than imaging controls and preHDs (all *p*-values<0.05). PreHDs also displayed atrophy relative to imaging controls (all *p*-values<0.05) except in the thalamus (*p*=0.99). There was no main effect of group on the volumes of the hippocampus and amygdala (all *p*-values>0.05).

EarlyHDs showed a cortical thinning in the left angular gyrus, the left occipital superior cortex, the right caudal part of the middle frontal cortex and the right lateral occipital lobe relatively to imaging controls (Fig. 3B, Table 3). EarlyHDs also had a thinner right lateral occipital cortex than preHDs (Fig. 3C, Table 3). Cortical thickness was similar in preHDs and imaging controls.

**Figure 3.**
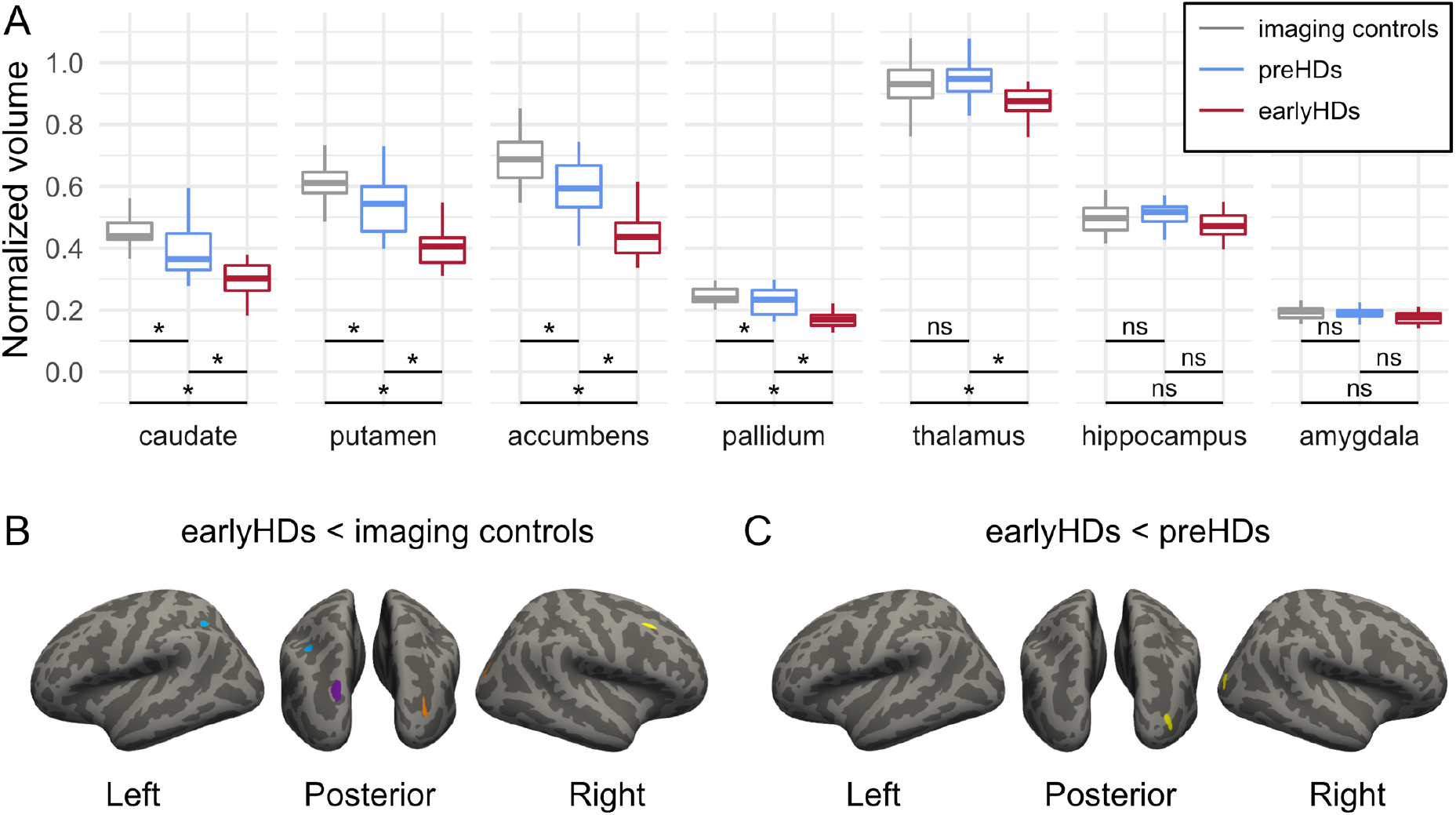
Neuroanatomical differences between groups. (A) Boxplots of the subcortical differences between groups. For representational purposes, volumes normalized by the total intracranial volume (tiv) are multiplied by 100 for all structures and by an additional 10 for the accumbens. In boxplots, the middle hinge corresponds to the median, the lower and upper hinges correspond respectively to the first and third quartiles. The whiskers extend from the hinge to the largest value no further than 1.5*inter-quartile range of the hinge. *: *p*<0.05, ns: non-significant. Imaging controls are represented in grey, premanifest participants (preHDs) in blue and Huntington’s disease patients at an early stage of the disease (earlyHDs) in red. (B) Cortical maps of differences between earlyHDs and imaging controls. Each cluster is represented in a different colour and identify a significant thinner cortex in earlyHDs compared to imaging controls. Blue: left angular gyrus, purple: left occipital superior, orange: right lateral occipital, yellow: right caudal middle frontal. (C) Cortical maps of differences between earlyHDs and controls. The cluster identifies a significant thinner cortex in earlyHDs compared to imaging controls. Yellow: right lateral occipital. In the cortical maps of panels (B) and (C), light grey represents gyrus and dark grey represents sulcus.

**Table 3.**
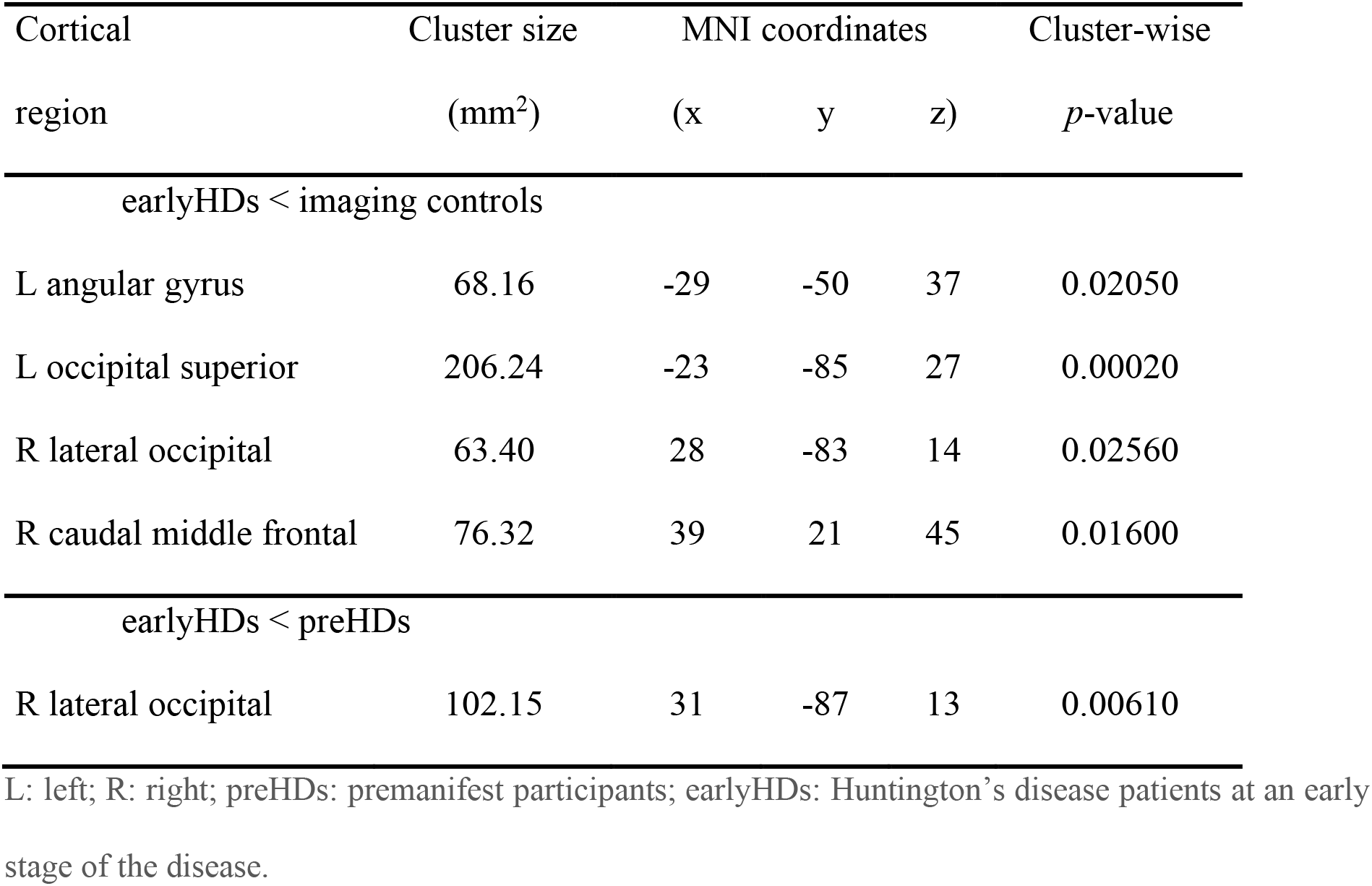
Cortical thickness differences between groups

#### Relationship between brain structure and behavioural measures in mutation carriers

##### Accuracy

Lower mean accuracies were predicted by lower accumbens volumes (*p*=0.005) independently of the diseases stage (no interaction, *p*=0.15). There was no significant cortical clusters associated with the mean accuracy (for one-tailed test see Supplementary Fig. 1A, Supplementary Table 2).

##### Reaction times

Slower mean reaction times were predicted by lower volumes of the accumbens, the putamen, the thalamus, the hippocampus and the pallidum (all *p*-values<0.05) independently of the disease stage (no interactions, all *p*-values>0.05)

Slower mean reaction times were also associated to a thinner cortex in clusters located in the left lateral and superior occipital cortex extending to the precuneus, and the right occipital cortex (all *p*-values<0.05), independently of the disease stage (no significant clusters with interaction, all *p*-values>0.05) (Supplementary Fig. 1B, Supplementary Table 2).

In all these significant cortical clusters, earlyHDs had a thinner cortex compared to imaging controls (all *p*-values<0.001) while there was no difference between preHDs and imaging controls (all *p*-values>0.05).

##### Drift rate

Lower drift rates were predicted by lower volumes of the accumbens, putamen and pallidum (all *p*-values<0.05) independently of the disease stage (no interactions, all *p*-values>0.05). In contrast, there was an interaction with the disease stage in the hippocampus (*t*(41)=-3.2, *p*=0.0026): a higher volume of the hippocampus predicted a higher drift rate in preHDs (*t*(41)=3.31, *p*=0.002) but not in earlyHDs (*t*(41)=-0.85, *p*=0.40) (Fig. 4A-B).

**Figure 4.**
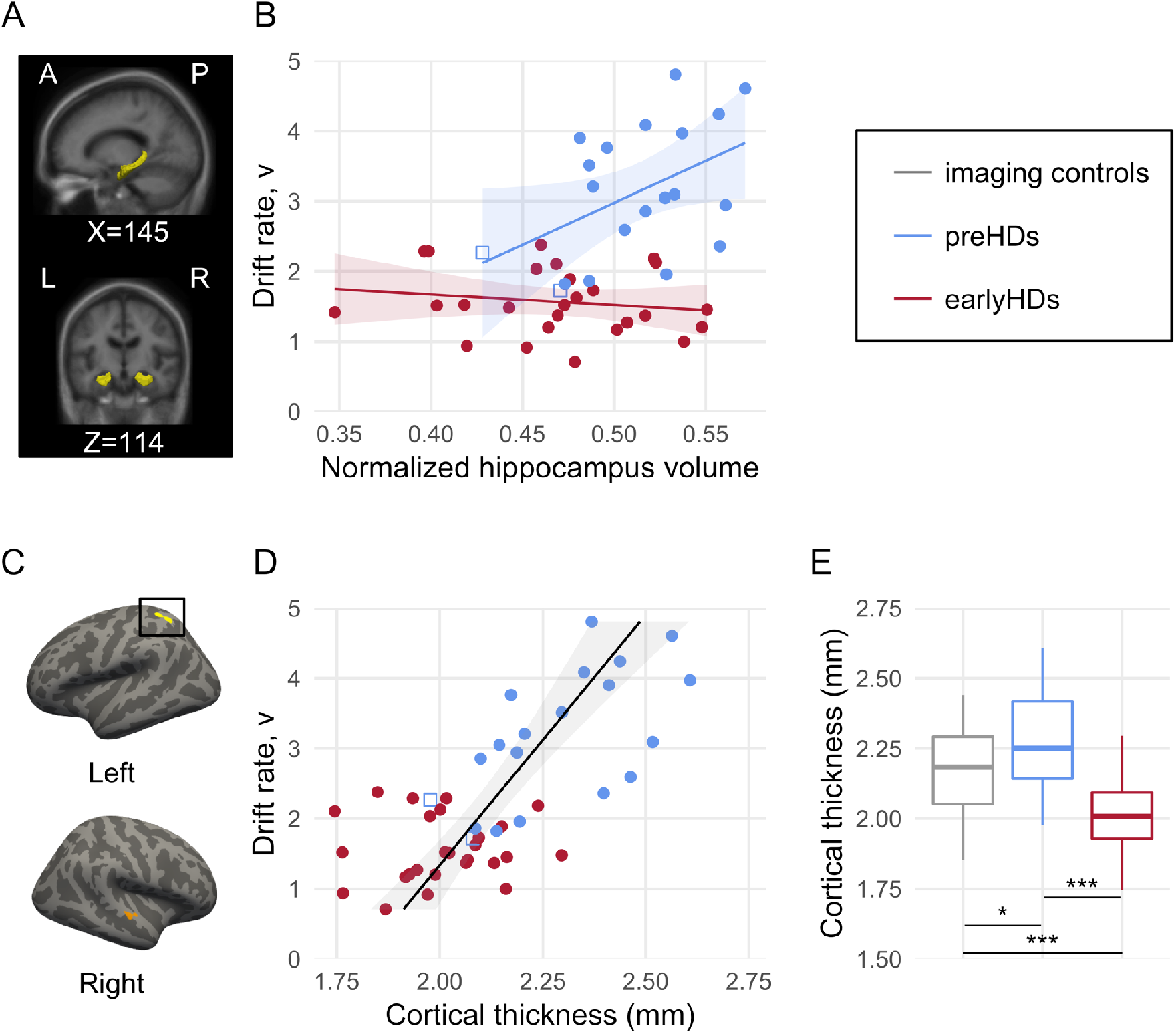
Relationship between brain structure and drift rate. (A) Hippocampus (yellow). X and Z are MNI coordinates of the sagittal and coronal views. A: anterior, P: posterior, R: right, L: left. (B) Relationship between the hippocampus volume normalized by the total intracranial volume (tiv) (*x*-axis) and the drift rate (*y*-axis). For representational purposes, the x-axis is multiplied by 100. (C) Cortical maps of significant clusters with a positive correlation between the cortical thickness and the drift rate. Light grey represents gyrus and dark grey represents sulcus. Yellow: left superior parietal cluster (152.71 mm^2^, MNI coordinates: [−34, −55, 59]); Orange: right superior temporal cluster (112.72 mm^2^, MNI coordinates: [63, −16, −1]). The black squares identify the cluster containing the data displayed in panels (D) and (E). (D) Relationship between the mean cortical thickness in the yellow cluster of interest (panel (C)) (*x*-axis) and the drift rate (*y*-axis). (E) Boxplots of the mean cortical thickness in the yellow cluster (panel (C)). In boxplots, the middle hinge corresponds to the median, the lower and upper hinges correspond relatively to the first and third quartiles. The whiskers extend from the hinge to the largest value no further than 1.5*inter-quartile range of the hinge. *: *p*<0.05, ***: *p*<0.001. Imaging controls are represented in grey, premanifest participants (preHDs) in blue and Huntington’s disease patients at an early stage of the disease (earlyHDs) in red. In (B) and (D), points represent individual values, lines and shades around them represent the linear fit and the confidence interval. The blue squares represent the two preHDs identified as closest to the disease onset.

Lower drift rates were also associated to a thinner cortex in clusters located in in the left superior parietal cortex and the right superior temporal gyrus (all *p*-values<0.05), independently of the disease stage (no significant clusters with interaction, all *p*-values>0.05) (Fig. 4C-D).

In these two significant cortical clusters, earlyHDs had a thinner cortex compared to imaging controls (all *p*-values<0.001) while there was no significant difference between preHDs and imaging controls (all *p*-values>0.05). Yet, preHDs tended to have a thicker cortex compared to imaging controls. Post-hoc, we identified two preHDs close to clinical onset with a negative estimated age from onset (Langbehn et al., 2004) (all other preHDs had a positive estimated age from onset) and a close to threshold total motor score of four (Tabrizi et al., 2009) (all other preHDs had a total motor score of one or zero). When we removed these two participants, we found that the preHDs had a thicker cortex in the left superior parietal cluster (*β*=0.12, 95%CI=[0.001,0.24], *p*<0.05) (Fig. 4E).

##### Response threshold

There was neither significant relationships between the neuroanatomical structure (subcortical and cortical) and the response threshold nor significant interaction with the disease stage (all *p*-values >0.05).

## Discussion

In this study, we aimed to identify the mechanism underlying cognitive compensation in neurodegenerative diseases using Huntington’s disease as a model. To address this issue, we used DDMs to gain insight into the decision processes involved in a language discrimination task (Fig. 1). As expected, analyses of behavioural performances based on accuracy and response times showed an absence of impairment in preHDs relative to controls (Fig. 2A-B). However, analyses of DDMs parameters highlighted a different profile in preHDs, with a nascent increase in response threshold predicting a faster drift rate of evidence accumulation (Fig. 2C-F). This faster rate of evidence accumulation presumably accounts for the observed compensation, by maintaining response times and accuracy at values similar to those of controls. In contrast, earlyHDs displayed impairment in both behavioural and DDMs analyses, suggesting that compensatory mechanisms were absent or insufficient to counterbalance the decline in these individuals. Both a higher response threshold and a slower rate of evidence accumulation resulted in lower accuracy and longer response times in this group (Fig. 2). As previously reported (Tabrizi et al., 2009), striatal atrophy was observed not only in earlyHDs but also in preHDs (Fig. 3A), while only earlyHDs presented a cortical thinning (Fig. 3). In preHDs, the spared thalamus (Fig. 3A), the relationship between the drift rate and both the volume of the hippocampus (Fig. 4B) and the superior parietal cortical hypertrophy, (Fig. 4E) suggest that the compensatory mechanism might be based on an increase in attention allocation.

A higher response threshold leads to more cautious and slower decision-making in perceptual decisions. In our task, the response threshold increased across disease stages suggesting a gradient of impairment during disease progression (Fig. 2C). Forstmann *et al*. (2010) showed that the flexible variation of the DDMs response threshold depended on the strength of the connections between the cortex and striatum inhibiting the subthalamic nucleus. The indirect pathway from the striatum to the thalamus through the external globus pallidus and subthalamic nucleus is more strongly affected in earlyHDs than in preHDs (André et al., 2010). Stimulation of the subthalamic nucleus in patients with Parkinson’s disease results in more impulsive choices and a decrease in response threshold (Cavanagh et al., 2011). According to the André *et al*. model (André et al., 2010), disruption of the indirect pathway and a decrease in the number of white matter fibres extending between the striatum and the cortex (Marrakchi-Kacem et al., 2013; Poudel et al., 2014) in Huntington’s disease increase inhibition of the subthalamic nucleus. This may account for the higher response threshold in mutation carriers and could explain why we did not find neuroanatomical correlates of the response threshold. Diffusion MRI data would be needed to further investigated this hypothesis.

This increase in response threshold might be expected to alter behavioural performances, as was observed in earlyHDs. However, the preHDs performed similarly to controls (Fig. 2A-B). We hypothesise that the higher drift rate in preHDs is a compensatory mechanism that preserves normal accuracy and response times (Fig. 2E). The association between the increase in response threshold and the increase of the drift rate of accumulation is consistent with compensation between these two processes (Fig. 2F). The stronger relationship in preHDs compared to earlyHDs and controls, together with the inability of earlyHDs to maintain normal behavioural performances, are consistent with models assuming that compensation mechanisms become less effective as the disease progresses and the pathological load increases, leading to observable cognitive impairments (Papoutsi et al., 2014; Gregory et al., 2018). Drift rate may therefore constitute a measurable cognitive marker of compensation.

Imaging seems appropriate to reveal neural compensation mechanisms. Indeed, active compensation mechanisms may depend on either an increase in the activity of a deficient network or the recruitment of alternative networks with available resources (Barulli and Stern, 2013). Both have been observed in preHDs, where functional imaging studies show changes in BOLD responses in task-dependent regions, despite similar behavioural performances to controls (Klöppel et al., 2009; Wolf et al., 2012). Although changes in connectivity, structure, or activation can provide information about the link between the disease and neural reorganisation, this link may be pathological rather than compensatory if not correlated to better performances (Papoutsi et al., 2014; Soloveva et al., 2018). Here, we observed left superior parietal cortex hypertrophy in preHDs (Fig. 4E). In addition, an increase in cortical thickness was associated with better performances (shorter response times, better accuracy, and higher drift rates) (Supplementary Fig. 1, Supplementary Table 2, Fig. 4C-D), strengthening the hypothesis of a successful compensation. Hyperactivation of the superior parietal cortex has been associated with motor compensatory mechanisms (Feigin et al., 2006). In a post-hoc analysis, the two preHDs closest to disease onset did not present this pattern of cortical thickening, further supporting the hypothesis of a compensatory mechanism failing as the pathological load increases. Previous studies on Alzheimer’s disease (Fortea et al., 2010) and Huntington’s disease (Nopoulos et al., 2010) have reported a preclinical stage of hypertrophy preceding atrophy in patients with symptoms. This may reflect an experience-dependent increase in neural volume (Luders et al., 2009; Suh et al., 2019) due to attempts to compensate for the dysregulation of brain regions and the striatum.

Drift rate is linked to attention in healthy subjects (Mulder et al., 2014) and patients with attention deficit and hyperactivity disorder (Ziegler et al., 2016). Individuals with higher attentional capacities accumulate evidence faster in a noisy perceptual decision task (Nunez et al., 2015). Here, independently of the disease stage, drift rate correlated positively with cortical thickness in the left superior parietal and the right superior temporal cortex – regions which are associated with a better ability to sustain attention (Fig. 4C-D) (Fan et al., 2005; Kristensen et al., 2013; Mitko et al., 2019). This suggests that mutation carriers with better attentional capacities have higher drift rates. Within this attentional network, the hippocampus plays a role in maintaining high-resolution representations in working memory when a complex and precise representation is required (Yonelinas, 2013), especially in online perception (Córdova et al., 2019). The content of working memory automatically modulates attention by gating the information matching its content into awareness (Soto et al., 2008). In preHDs, a larger hippocampal volume predicted a higher drift rate (Fig. 4B). This raises the possibility that preHDs use the hippocampus to tune their attention to relevant stimulus features (fined-grained representation of pseudowords in our task), increasing information extraction and leading to faster rate of evidence accumulation. The structural modification (hypertrophy) (Fig. 4E) observed in preHDs suggests a daily-used mechanism not limited to our task. In contrast, hippocampal volume is not related to drift rate in earlyHDs. This suggests that attentional tuning allowing for drift rate adjustment is no longer efficient as the disease proceeds. The inability of earlyHDs to recruit sufficient additional attentional resources is consistent with their brain atrophy and the pattern of attention impairment observed in this disease. In our cohort, both the right caudal part of the middle frontal cortex and the thalamus are atrophied in earlyHDs, but spared in preHDs (Fig. 3). These structures are key components of the attentional network (Fan et al., 2005; Corbetta et al., 2008). The literature reports that preHDs are minimally affected in this domain, whereas earlyHDs present a wide range of attentional deficits (e.g. sustained attention (Hart et al., 2012)). The atrophy of earlyHDs in the left angular gyrus, a key structure involved in phonological discrimination (Jacquemot et al., 2003), has been previously reported (Macdonald et al., 1997). This might also prevent them from biasing their attention to fine grained phonological features present in our task.

Despite this study’s small cohort size, DDMs detected differences between preHDs and controls and identified cognitive processes that may underlie compensation mechanisms in a language discrimination task. This demonstrates the added value of DDMs, combined with language relatively, to classical neuropsychological tests, which rarely detect differences in preHDs, other than in cohorts of hundreds of participants (Paulsen et al., 2004; Tabrizi et al., 2009). Here, the use of a linguistic task was motivated by studies showing the sensitivity of language in detecting subtle disorders in small cohorts of preHDs (de Diego Balaguer et al., 2008; Németh et al., 2012; Hinzen et al., 2018). The duration of the task and its simplicity (pseudoword discrimination) make it easily adaptable and transferable to other languages. Showing the generality of the drift rate as a marker of compensation, and of attention allocation increase as a compensatory mechanism in preHDs would require assessing other cognitive domains. Structural imaging provides a hint into the attentional network. Yet, studying the functional correlates of the drift rate in Huntington’s disease would provide a greater understanding of online allocation of attentional resources as a compensatory mechanism. Focusing on the role of the superior parietal cortex and hippocampus seems promising in Huntington’s disease but would need to be replicated in other neurodegenerative diseases and studied over time.

## Data Availability

Participants signed an informed consent form guaranteeing data confidentiality, and we were therefore unable to share our data through deposition in an open-access repository. However, the data are available, on request, from Prof. Anne-Catherine Bachoud-Levi, for research purposes only. The person requesting the data must sign a confidentiality agreement provided by Assistance Publique-Hopitaux de Paris stipulating that they will make no attempt to identify participants. Given the small size of the cohorts studied, the identification of mutation carriers from center information is feasible. Thus, even with a signed confidentiality agreement, some of the information will be removed, to ensure confidentiality.

## Conflict of interest

The authors declare no competing financial interests.

## Acknowledgments

This work was supported by the Agence Nationale de la Recherche (**ANR-17-EURE-0017**, and **ANR-11-JSH2-006-1**), a grant from *Fondation Maladies Rares, programme Sciences Humaines et Sociales & Maladies Rares* awarded to Charlotte Jacquemot, a PhD grant from *Ecole Doctorale Sciences de la vie et de la santé* ED402 awarded to Lorna Le Stanc and an interface contract (Institut National de la Santé Et de la Recherche Médicale - INSERM) awarded to Anne-Catherine Bachoud-Lévi. The Henri Mondor Hospital National Reference Centre for Huntington’s Disease funded the follow-up of all the patients included in this study (Ministry of Health). We thank Laurent Cleret de Langavant for his insight concerning the imaging results, Lucas Filipin for assistance with the software, and Renaud Massart and Karen Hernandez for proofreading the manuscript.

**Abbreviations**
DDMs: = Drift Diffusion Models
earlyHDs: = Huntington’s disease patients at an early stage of the disease
preHDs: = participants with premanifest Huntington’s disease

## Notes

### Competing Interest Statement

The authors have declared no competing interest.

### Author Declarations

Participants were recruited from a clinical biomarker study (NCT01412125) in outpatients approved by the ethics committee of Henri Mondor Hospital (Creteil, France)

## References

Anders R, Riès S, Van Maanen L, Alario FX (2017) Lesions to the left lateral prefrontal cortex impair decision threshold adjustment for lexical selection. Cogn Neuropsychol 34:1–20.

André VM, Cepeda C, Levine MS (2010) Dopamine and Glutamate in Huntington’s Disease: A Balancing Act. CNS Neurosci Ther 16:163–178.

Andrews SC, Domínguez JF, Mercieca E-C, Georgiou-Karistianis N, Stout JC (2015) Cognitive interventions to enhance neural compensation in Huntington’s disease. Neurodegener Dis Manag 5:155–164 Available at: http://www.futuremedicine.com/doi/10.2217/nmt.14.58.

Barulli D, Stern Y (2013) Efficiency, capacity, compensation, maintenance, plasticity: Emerging concepts in cognitive reserve. Trends Cogn Sci 17:502–509 Available at: http://dx.doi.org/10.1016/j.tics.2013.08.012.

Cavanagh JF, Wiecki T V, Cohen MX, Figueroa CM, Samanta J, Sherman SJ, Frank MJ (2011) Subthalamic nucleus stimulation reverses mediofrontal influence over decision threshold. Nat Neurosci 14:1462–1467 Available at: http://www.nature.com/doifinder/10.1038/nn.2925.

Corbetta M, Patel G, Shulman GL (2008) The Reorienting System of the Human Brain: From Environment to Theory of Mind. Neuron 58:306–324.

Córdova NI, Turk_Browne NB, Aly M (2019) Focusing on what matters: Modulation of the human hippocampus by relational attention. Hippocampus:1–13.

de Diego Balaguer R, Couette M, Dolbeau G, Dürr A, Youssov K, Bachoud-Lévi A-C (2008) Striatal degeneration impairs language learning: evidence from Huntington’s disease. Brain 131:2870–2881.

Dehaene-Lambertz G, Baillet S (1998) A phonological representation in the infant brain. Neuroreport 9:1885–1888.

Fan J, McCandliss BD, Fossella J, Flombaum JI, Posner MI (2005) The activation of attentional networks. Neuroimage 26:471–479.

Feigin A, Ghilardi MF, Huang C, Ma Y, Carbon M, Guttman M, Paulsen JS, Ghez CP, Eidelberg D (2006) Preclinical Huntington’s disease: Compensatory brain responses during learning. Ann Neurol 59:53–59.

Fischl B, Dale AM (2000) Measuring the thickness of the human cerebral cortex from magnetic resonance images. Proc Natl Acad Sci U S A 97:11050–11055 Available at: http://www.pnas.org/cgi/doi/10.1073/pnas.200033797.

Fischl B, Salat DH, Busa E, Albert M, Dieterich M, Haselgrove C, Kouwe A Van Der, Killiany R, Kennedy D, Klaveness S, Montillo A, Makris N, Rosen B, Dale AM (2002) Neurotechnique Whole Brain Segmentation: Automated Labeling of Neuroanatomical Structures in the Human Brain. Neuron 33:341–355 Available at: https://ac.els-cdn.com/S089662730200569X/1-s2.0-S089662730200569X-main.pdf?_tid=;6c258edb-4f39-433a-9bed-64f82c122596&acdnat=;1547096266_d8dd0eae210187d84e43084ef94991df.

Fischl B, Sereno M, Dale A (1999) Cortical surface-based analysis. Neuroimage 9:195–207 Available at: http://citeseerx.ist.psu.edu/viewdoc/download?doi=;10.1.1.15.6598&rep=;rep1&type=;pdf.

Forstmann BU, Anwander A, Schäfer A, Neumann J, Brown S, Wagenmakers E-J, Bogacz R, Turner R (2010) Cortico-striatal connections predict control over speed and accuracy in perceptual decision making. Proc Natl Acad Sci U S A 107:15916–15920 Available at: http://www.pnas.org/content/107/36/15916.abstract.

Fortea J, Sala-Llonch R, Bartrés-Faz D, Bosch B, Lladó A, Bargalló N, Molinuevo JL, Sánchez-Valle R (2010) Increased cortical thickness and caudate volume precede atrophy in psen1 mutation carriers. J Alzheimer’s Dis 22:909–922.

Goodrich B, Gabry J, Ali I, Brilleman S (2020) rstanarm: Bayesian applied regression modeling via Stan. R package version 2.19.3. Available at: https://mc-stan.org/rstanarm.

Gregory S et al. (2018) Testing a longitudinal compensation model in premanifest Huntington’s disease. Brain 141:2156–2166.

Hart EP, Dumas EM, Reijntjes RHAM, Van Der Hiele K, Van Den Bogaard SJA, Middelkoop HAM, Roos RAC, Van Dijk JG (2012) Deficient sustained attention to response task and P300 characteristics in early Huntington’s disease. J Neurol 259:1191–1198.

Hinzen W, Rosselló J, Morey C, Camara E, Garcia-Gorro C, Salvador R, de Diego Balaguer R (2018) A systematic linguistic profile of spontaneous narrative speech in pre-symptomatic and early stage Huntington’s disease. Cortex 100:71–83.

Huntington Study Group (1996) Unified Huntington’s Disease Rating Scale: Reliability and Consistency. Mov Disord 11:136–142.

Jacquemot C, Pallier C, LeBihan D, Dehaene S, Dupoux E (2003) Phonological grammar shapes the auditory cortex: a functional magnetic resonance imaging study. J Neurosci 23:9541–9546 Available at: http://www.ncbi.nlm.nih.gov/pubmed/14573533.

Klöppel S, Draganski B, Siebner HR, Tabrizi SJ, Weiller C, Frackowiak RSJ (2009) Functional compensation of motor function in pre-symptomatic Huntingtons disease. Brain 132:1624–1632.

Kristensen LB, Wang L, Petersson KM, Hagoort P (2013) The interface between language and attention: Prosodic focus marking recruits a general attention network in spoken language comprehension. Cereb Cortex 23:1836–1848.

Langbehn DR, Brinkman RR, Falush D, Paulsen JS, Hayden MR (2004) A new model for prediction of the age of onset and penetrance for Huntington’s disease based on CAG length. Clin Genet 65:267–277.

Luders E, Toga AW, Lepore N, Gaser C (2009) The underlying anatomical correlates of long-term meditation: Larger hippocampal and frontal volumes of gray matter. Neuroimage 45:672–678 Available at: http://dx.doi.org/10.1016/j.neuroimage.2008.12.061.

Macdonald V, Halliday GM, Trent RJ, McCusker EA (1997) Significant loss of pyramidal neurons in the angular gyrus of patients with Huntington’s disease. Neuropathol Appl Neurobiol 23:492–495.

Malejko K, Weydt P, SüßBmuth SD, Grön G, Landwehrmeyer BG, Abler B (2014) Prodromal Huntington disease as a model for functional compensation of early neurodegeneration. PLoS One 9:1–14.

Marrakchi-Kacem L, Delmaire C, Guevara P, Poupon F, Lecomte S, Tucholka A, Roca P, Yelnik J, Durr A, Mangin JF, Lehéricy S, Poupon C (2013) Mapping Cortico-Striatal Connectivity onto the Cortical Surface: A New Tractography-Based Approach to Study Huntington Disease. PLoS One 8:e53135.

Mattis S (1976) Mental status examination for organic mental syndrome in elderly patients. In: Geriatric Psychiatry (Bellak L, Karasu TB, eds), pp 77–121. New-York: Grune & Straton.

Mitko A, Rothlein D, Poole V, Robinson M, Mcglinchey R, Degutis J, Salat D, Esterman M (2019) Individual differences in sustained attention are associated with cortical thickness. Hum Brain Mapp: 1–11.

Mulder MJ, van Maanen L, Forstmann BU (2014) Perceptual decision neurosciences - a model-based review. Neuroscience 277:872–884 Available at: http://dx.doi.org/10.1016/j.neuroscience.2014.07.031.

Näätänen R, Lehtokoski A, Lennes M, Cheour M, Huotilainen M, Iivonen A, Vainio M, Alku P, Ilmoniemi RJ, Luuk A, Allik J, Sinkkonen J, Alho K (1997) Language-specific phoneme representations revealed by electric and magnetic brain responses. Nature 385:432–434.

Németh D, Dye CD, Sefcsik T, Janacsek K, Turi Z, Londe Z, Klivenyi P, Kincses TZ, Nikoletta S, Vecsei L, Ullman MT (2012) Language deficits in Pre-Symptomatic Huntington’s Disease: Evidence from Hungarian. Brain Lang 121:248–253.

Nopoulos PC, Aylward EH, Ross CA, Johnson HJ, Magnotta VA, Juhl AR, Pierson RK, Mills J, Langbehn DR, Paulsen JS (2010) Cerebral cortex structure in prodromal Huntington disease. Neurobiol Dis 40:544–554 Available at: http://dx.doi.org/10.1016Zj.nbd.2010.07.014.

Nunez MD, Srinivasan R, Vandekerckhove J (2015) Individual differences in attention influence perceptual decision making. Front Psychol 8:1–13.

Papoutsi M, Labuschagne I, Tabrizi SJ, Stout JC (2014) The cognitive burden in Huntington’s disease: Pathology, phenotype, and mechanisms of compensation. Mov Disord 29:673–683.

Paulsen JS, Zimbelman JL, Hinton SC, Langbehn DR, Leveroni CL, Benjamin ML, Reynolds NC, Rao SM (2004) fMRI biomarker of early neuronal dysfunction in presymptomatic Huntington’s disease. Am J Neuroradiol 25:1715–1721.

Pini L, Jacquemot C, Cagnin A, Meneghello F, Semenza C, Mantini D, Vallesi A (2019) Aberrant brain network connectivity in presymptomatic and manifest Huntington’s disease: A systematic review. Hum Brain Mapp:1–14.

Poudel GR, Stout JC, Domínguez D JF, Salmon L, Churchyard A, Chua P, Georgiou-Karistianis N, Egan GF (2014) White matter connectivity reflects clinical and cognitive status in Huntington’s disease. Neurobiol Dis 65:180–187 Available at: http://dx.doi.org/10.1016/j.nbd.2014.01.013.

Ratcliff R, Childers R (2015) Individual differences and fitting methods for the two-choice diffusion model of decision making. Decision 2:237–279.

Ratcliff R, Mckoon G (2008) The Diffusion Decision Model: Theory and Data for Two-Choice Decision Tasks. Neural Comput 20:873–922.

Ross CA, Reilmann R, Cardoso F, McCusker EA, Testa CM, Stout JC, Leavitt BR, Pei Z, Landwehrmeyer B, Martinez A, Levey J, Srajer T, Bang J, Tabrizi SJ (2019) Movement Disorder Society Task Force Viewpoint: Huntington’s Disease Diagnostic Categories. Mov Disord Clin Pract 6:541–546.

Scheller E, Abdulkadir A, Peter J, Tabrizi SJ, Frackowiak RSJ, Klöppel S (2013) Interregional compensatory mechanisms of motor functioning in progressing preclinical neurodegeneration. Neuroimage 75:146–154 Available at: http://dx.doi.org/10.1016/j.neuroimage.2013.02.058.

Snowden JS, Craufurd D, Thompson J, Neary D (2002) Psychomotor, Executive, and Memory Function in Preclinical Huntington’s Disease. J Clin Exp Neuropsychol 24:133–145 Available at: http://www.tandfonline.com/doi/abs/10.1076/jcen.24.2.133.998.

Soloveva M V., Jamadar SD, Poudel G, Georgiou-Karistianis N (2018) A critical review of brain and cognitive reserve in Huntington’s disease. Neurosci Biobehav Rev 88:155–169 Available at: https://doi.org/10.1016/j.neubiorev.2018.03.003.

Soto D, Hodsoll J, Rotshtein P, Humphreys GW (2008) Automatic guidance of attention from working memory. Trends Cogn Sci 12:342–348.

Stout JC, Jones R, Labuschagne I, O’Regan AM, Say MJ, Dumas EM, Queller S, Justo D, Dar Santos R, Coleman A, Hart EP, Dürr A, Leavitt BR, Roos RA, Langbehn DR, Tabrizi SJ, Frost C (2012) Evaluation of longitudinal 12 and 24 month cognitive outcomes in premanifest and early Huntington’s disease. J Neurol Neurosurg Psychiatry 83:687–694.

Suh JS, Schneider MA, Minuzzi L, Glenda M, Strother SC, Kennedy SH, Benicio N (2019) Cortical thickness in major depressive disorder: a systematic review and meta-analysis. Prog Neuropsychopharmacol Biol Psychiatry 88:287–302.

Tabrizi SJ, Langbehn DR, Leavitt BR, Roos RA, Durr A, Craufurd D, Kennard C, Hicks SL, Fox NC, Scahill RI, Borowsky B, Tobin AJ, Rosas HD, Johnson H, Reilmann R, Landwehrmeyer GB, Stout JC (2009) Biological and clinical manifestations of Huntington’s disease in the longitudinal TRACK-HD study: cross-sectional analysis of baseline data. Lancet Neurol 8:791–801 Available at: http://linkinghub.elsevier.com/retrieve/pii/S147444220970170X.

Tabrizi SJ, Scahill RI, Durr A, Roos RA, Leavitt BR, Jones R, Landwehrmeyer GB, Fox NC, Johnson H, Hicks SL, Kennard C, Craufurd D, Frost C, Langbehn DR, Reilmann R, Stout JC (2011) Biological and clinical changes in premanifest and early stage Huntington’s disease in the TRACK-HD study: The 12-month longitudinal analysis. Lancet Neurol 10:31–42 Available at: http://dx.doi.org/10.1016/S1474-4422(10)70276-3.

Wiecki T V., Antoniades CA, Stevenson A, Kennard C, Borowsky B, Owen G, Leavitt B, Roos R, Durr A, Tabrizi SJ, Frank MJ (2016) A computational cognitive biomarker for early-stage Huntington’s disease. PLoS One 11:e0148409.

Wiecki T V, Sofer I, Michael FJ (2013) HDDM: Hierarchical Bayesian estimation of the Drift-Diffusion Model in Python. Front Neuroinform 7:1–10.

Wolf RC, Gron G, Sambataro F, Vasic N, Wolf ND, Thomann PA, Saft C, Landwehrmeyer GB, Orth M (2012) Brain activation and functional connectivity in premanifest Huntington’s disease during states of intrinsic and phasic alertness. Hum Brain Mapp 33:2161–2173.

Yonelinas AP (2013) The hippocampus supports high-resolution binding in the service of perception, working memory and long-term memory. Behav Brain Res 254:34–44 Available at: http://dx.doi.org/10.1016/j.bbr.2013.05.030.

Zhang J, Rittman T, Nombela C, Fois A, Coyle-Gilchrist I, Barker RA, Hughes LE, Rowe JB (2016) Different decision deficits impair response inhibition in progressive supranuclear palsy and Parkinson’s disease. Brain 139:161–173.

Ziegler S, Pedersen ML, Mowinckel AM, Biele G (2016) Modelling ADHD: A review of ADHD theories through their predictions for computational models of decision-making and reinforcement learning. Neurosci Biobehav Rev 71:633–656 Available at:http://dx.doi.org/10.1016/j.neubiorev.2016.09.002.

